# The Impact of Race, Ethnicity, and Socioeconomic Status on the Severity of Menopause Symptoms: A Study of 68,864 Women

**DOI:** 10.1101/2023.12.21.23299398

**Authors:** Alison Kochersberger, Aeowynn Coakley, Leah Millheiser, Jerrine R. Morris, Claire Manneh, Alicia Jackson, Jennifer L. Garrison, Eduardo Hariton

## Abstract

**Objective:** To evaluate how race, ethnicity, and socioeconomic status (SES) affect the severity of menopause symptoms in a large, diverse sample of women.

**Methods:** For this cross-sectional study conducted between March 24, 2019, and January 13, 2023, a total of 68,864 women were enrolled from the Evernow online telehealth platform. Participants underwent a clinical intake survey, which encompassed demographic information, detailed medical questionnaires, and a modified Menopause Rating Scale (MRS). The modified scale was adapted for ease of use online and is available in the supplementary material along with the full intake. Symptom severity was evaluated using a multivariate binomial generalized linear model, accounting for factors such as race, ethnicity, age, BMI, smoking status, bilateral oophorectomy status, and SES. Odds ratios and confidence intervals were calculated based on the linear regression coefficients.

**Results:** Of the participants, 67,867 (98.6%) were included in the analysis after excluding outliers and those with unknown oophorectomy status. The majority of respondents identified as White (77.4%), followed by Hispanic (9.0%), Black (6.7%), two or more races/ethnicities (4.4%), Asian (1.2%), Indigenous/First Nations (0.8%), Middle Eastern (0.3%), and South Asian (0.2%). Notably, individuals identifying as Black, Hispanic, Indigenous/First Nations, Middle Eastern, or with two or more races/ethnicities reported higher levels of symptom severity compared to their White counterparts. Conversely, Asian and South Asian participants reported lower symptom severity. Even after incorporating SES into the linear model, racial and ethnic groups with lower SES (Black, Hispanic, Indigenous, and multiple ethnicities) exhibited slight shifts in odds ratios, while maintaining high statistical significance.

**Conclusions:** Our study suggests that the relationship between race and ethnicity and the severity of menopause symptoms is not solely explained by differences in SES but is itself an independent factor. Understanding and addressing social, cultural, and economic factors is crucial to reduce disparities in menopausal symptoms.

## INTRODUCTION

Menopause is a biological process that occurs in women and gender-diverse individuals as they age, characterized by the cessation of menstrual periods and a decline in hormone production. These hormonal changes are associated with various physical and psychological symptoms, with vasomotor symptoms lasting a median of 7.4 years and having a profound impact on a woman’s quality of life^1–4^. Race and ethnicity, as well as socioeconomic status, may influence the experience of menopause symptoms.

The Study of Women’s Health Across the Nation (SWAN) is a multiethnic, longitudinal, cohort study of mid-life women. This was the first NIH observational study of its kind and included women from different backgrounds to characterize the menopausal experience of a diverse group of women^5^. SWAN found that vasomotor symptoms were more prevalent in African-American and Hispanic women, while vaginal dryness was most prevalent in Hispanic women^6^. Another analysis of the SWAN study found that low-income women reported greater severity and frequency of menopausal symptoms compared to White women and women with higher income; however, these findings should be interpreted with caution, as they can be influenced by factors such as access to healthcare and use of hormone therapy, which varies based on demographic characteristics^7,8^. Other studies have sought to describe specific symptoms and their relationship to race and ethnicity, but none of them examined all symptoms across the Menopause Rating Scale (MRS)^9–13^. Further research is needed to explore the relationship between race and ethnicity, socioeconomic status, and menopausal symptoms.

The objective of the current study is to evaluate how race, ethnicity, and socioeconomic status affect the severity of menopause symptoms in a large, diverse sample of women, expanding on prior research by including granularity in self-reported racial and ethnic groups, and evaluating socioeconomic status as a covariate. The aim is to shed light on the complex role that these factors play in menopausal symptomatology to better inform treatment interventions to improve the quality of life of menopausal women.

## METHODS

### Survey Development

Evernow is a telehealth platform that provides evidence-based menopause care. Prospective patients must complete a clinical intake survey, which includes demographic information, a medical history questionnaire, and a modified Menopause Rating Scale to determine eligibility and screen for contraindications. The survey questions are close-ended with answer options provided to participants. Survey question details and answer choices are in the Supplemental Methods.

### Participant Eligibility and Recruitment

Participants signed up for care at Evernow, advertised through digital marketing and via word of mouth. Patients usually pay out of pocket for treatment, which varied by the medication recommended by the provider, as well as during the time of the study. Only participants living in the United States were eligible to participate. A total of 68,864 participants were administered a clinical intake survey from March 24, 2019 to January 13, 2023. The survey was administered to prospective participants who self-identified as experiencing one or more symptoms common to perimenopause or menopause. All Evernow prospective patients consented to having their data de-identified and aggregated for the purposes of research. This study was reviewed by the Institutional Review Board (IRB) of the Buck Institute for Research on Aging and was deemed exempt from needing IRB approval.

### Survey Analysis

Survey results were de-identified before the analysis. For symptom severity, a rating of 1 or 2 was classified as “not severe”, and a rating of 3 or 4 was classified as “severe”. Participants who did not experience a given symptom were removed from severity analysis for that symptom. Participants identifying as East Asian, Asian American, or Southeast Asian were categorized as Asian. Those identifying as African-American or African were categorized as Black. Any participant who identified as more than one race or ethnicity were assigned to the Two or More Ethnicities category. Age was separated into those younger than 52 and those 52 or older. Smoking status combined those who actively used tobacco via cigarettes, vapes, or other. BMI was stratified by CDC BMI indices of underweight (<18.5), healthy weight (18.5-24.9), overweight (25.0-29.5, obesity (30.0-39.9), and severe obesity (BMI 40.0-59.9). Neighborhood affluence and disadvantage scores were stratified into four groups corresponding to quartiles within our dataset. Outliers for age (>70) and BMI (<15 and ≥60) were excluded from analysis. Additionally, those who reported a history of a hysterectomy but oophorectomy status was unknown were excluded.

The study used the National Neighborhood Data Archive (NaNDA) dataset to incorporate socioeconomic status into the analysis. Affluence and disadvantage scores were used as proxies for socioeconomic status, and were reflective of the time period from 2013-2017. Affluence score reflects the proportion of households with income greater than $75K, proportion of age 16+ employed in professional or managerial occupations, and proportion of adults with a Bachelor’s Degree or higher. Disadvantage score incorporates the proportion of female headed families with children, households with public assistance income or food stamps, families with income below the federal poverty level, and population age 16+ who are unemployed. ZIP codes of survey participants were crosswalked to ZIP Code Tabulation Areas (ZCTAs) using UDS Mapper’s 2017 crosswalk data, and affluence and disadvantage scores were assigned to survey participants.

Prevalence of severe symptoms for each variable was calculated and tested for multicollinearity before inclusion in the model. Multivariate binomial generalized linear models were created with symptom severity, affluence score and disadvantage score as dependent variables and race and ethnicity, affluence score and disadvantage score as the independent variables. Potential confounders that were controlled for in adjusted models included age, BMI, smoking status, bilateral oophorectomy status, affluence, disadvantage scores, and all hysterectomies for unusual periods. White was used as a reference for race and ethnicity. Odds ratios, confidence intervals, and p-values were calculated from the linear regression coefficients. All analyses were performed in R.

## RESULTS

In this study, 68,864 participants completed an online survey. Outliers for BMI, age and participants with unknown oophorectomy status were excluded, leaving 67,867 participants for analysis. The majority identified as White (77.4%), followed by Hispanic (9.0%), Black (6.7%), two or more races/ethnicities (4.4%), Asian (1.2%), Indigenous/First Nations (0.8%), Middle Eastern (0.3%), and South Asian (0.2%) (Table 1). About 45% of participants were under the age of 52, and the majority were overweight or obese (37.9% and 36.5%, respectively). Thirteen percent of participants were smokers. Of the 29.7% of participants who had hysterectomies, 15.8% did not have their ovaries removed, 2.3% reported unilateral oophorectomies, and 11.6% reported bilateral oophorectomies.

**TABLE 1.** Demographics.

| Characteristic | No. (%) (N=67867) |
| --- | --- |
| **Race/Ethnicity** |  |
| Asian | 790 (1.2) |
| Black | 4579 (6.7) |
| Hispanic/Latinx | 6116 (9.0) |
| Indigenous/First Nations | 531 (0.8) |
| Middle Eastern | 206 (0.3) |
| South Asian | 161 (0.2) |
| Two or More Ethnicities | 2970 (4.4) |
| White | 52514 (77.4) |
| **Age** |  |
| <52 | 30568 (45.0) |
| ≥52 | 37299 (55.0) |
| **BMI** |  |
| <18.5 | 424 (0.6) |
| 18.5-24.9 | 15137 (22.3) |
| 25-29.9 | 23227 (34.2) |
| 30-39.9 | 24267 (35.8) |
| ≥40 | 4812 (7.1) |
| **Smoking** |  |
| Non-Smoker | 59011 (87.0) |
| Smoker | 8856 (13.0) |
| **Hysterectomy** |  |
| None | 47734 (70.3) |
| Uterus | 10702 (15.8) |
| Unilateral |  |
| Oophorectomy | 1582 (2.3) |
| Bilateral Oophorectomy | 7849 (11.6) |

For our analysis we determined the odds ratios of severe symptoms (score 3 or 4), for each racial and ethnic group, using White as the reference. Within this analysis participants identifying as Black, Hispanic, Indigenous/First Nations, Middle Eastern, and two or more races/ethnicities, reported greater symptom severity as compared to those identifying as White for many symptoms (Fig. 1, Table 2, and Supplemental Table 1). For almost all symptoms, participants identifying as Asian or South Asian were found to have similar symptom severity than White participants. The symptom severity profile, however, varied across races and ethnicities. Those identifying as Black had the highest odds ratios for severe vasomotor symptoms (hot flashes (OR=1.91, 97.5% CI[1.76,2.09]), night sweats (OR=1.87, 97.5% CI[1.71,2.04]), sleep disturbances (OR= 1.67, 97.5% CI[1.51,1.86]), Table 2). Unusual periods (OR=1.66, 97.5% CI[1.37,2.04]), skin/hair changes (OR=1.65, 97.5% CI[1.49,1.84]), increased facial hair/acne (OR=1.48, 97.5% CI[1.48,1.31]), heart discomfort (OR=1.59, 97.5% CI[1.32,1.78]), and mood swings (OR=1.53, 97.5% CI[1.39,1.68]) were also the most severe for Black women. When BMI is accounted for, weight changes are least severe for Black women (OR=0.92, 97.5% CI[0.81,1.03]). Indigenous/First Nations participants experienced the most severe anxiety/depression (OR=1.70, 97.5% CI[1.31,2.24]), painful sex (OR=1.77, 97.5% CI[1.19,2.74]), and joint/muscular discomfort (OR=1.73, 97.5% CI[1.29,2.36]).

**FIG. 1.**
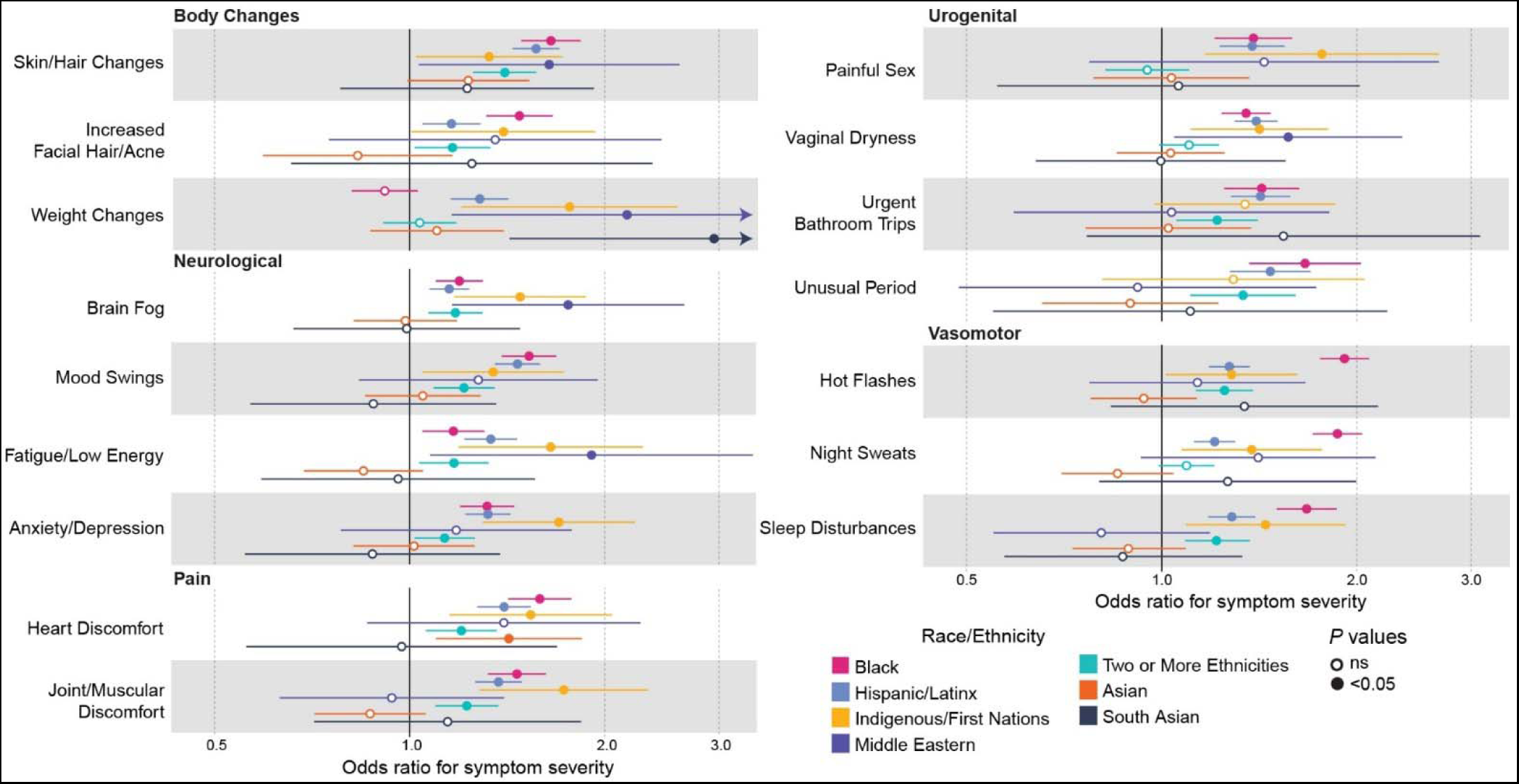
Adjusted Odds Ratios for Symptom Severity by Race/Ethnicity. Adjusted for age, BMI, smoking, and oophorectomy status. For unusual periods adjusted for all hysterectomy. Reference = White. Symptom Scores: 1-2 = Not Severe, 3-4 = Severe. Arrows indicate 95% CI extends beyond range.

**TABLE 2.**
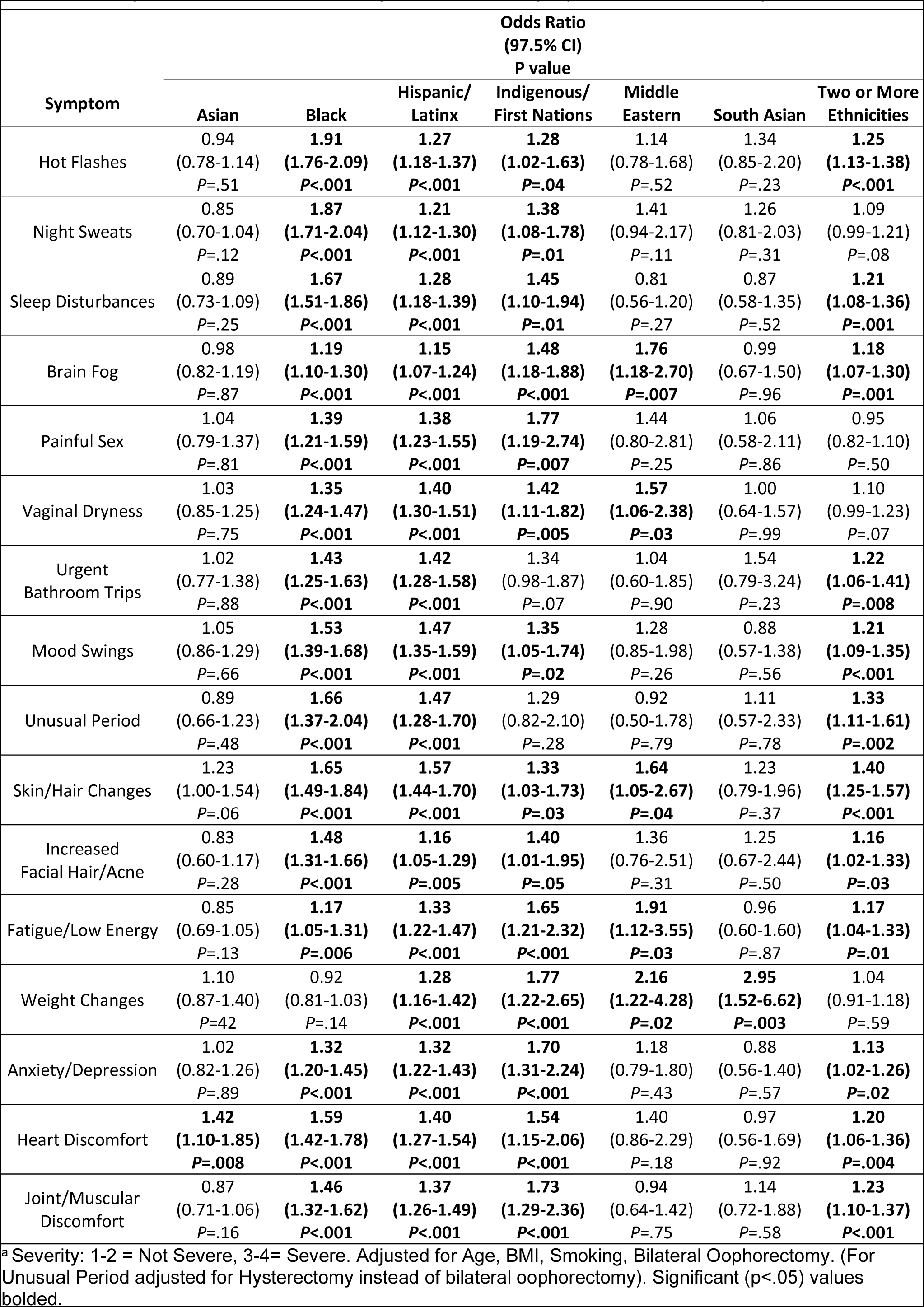
Adjusted Odds Ratios for Symptom Severity by Race and Ethnicity^a^.

Middle Eastern participants had the highest severity for brain fog (OR=1.76, 97.5% CI[1.18,2.70]) fatigue/low energy (OR=1.91, 97.5% CI[1.12,3.55]), and vaginal dryness (OR=1.57, 97.5% CI[1.06,2.38]). Hispanic identifying women have more severe symptoms than White women, with all symptoms having statistically significant (p<0.01) odds ratios greater than 1.16. Those who identified as two or more races/ethnicities have slightly more severe symptoms than those identifying as White, except for painful sex which was slightly less severe and not significant (OR=0.95, 97.5% CI[0.82,1.10]) and night sweats, vaginal dryness, and weight changes which were more severe but not significant. Of note, Asian women appear to experience significantly worse heart discomfort (OR=1.42, 97.5% CI[1.10,1.85]) and South Asian respondents had the highest severity for weight changes of all races and ethnicities (OR=2.95, 97.5% CI[1.52,6.62]). When calculating the prevalence of symptoms by either presence or absence, we found that elevated symptom severity did not always correspond with greater symptom frequency (Supplemental Table 2).

As race and ethnicity are social constructs, we investigated the impact of socioeconomic status (SES) on symptom severity. We used the affluence and disadvantage scores for the ZCTA corresponding to each respondent as a proxy for their SES. Participants in our survey have a similar SES compared to the general population. The mean affluence score for survey participants is 0.394 and the United States population is 0.326 (Supplemental Table 3). For disadvantage scores, survey participants have a mean of 0.096, while the general population is 0.099. We stratified affluence and disadvantage scores into four categories, with 1 being the lowest and 4 the highest. Across all symptoms, lower affluence score corresponded to higher symptom severity, with varying magnitudes of impact for each symptom (Supplemental Fig. 4 and Supplemental Table 4). The results when analyzing symptom severity by disadvantage score were similar (Supplemental Fig. 2 and Supplemental Table 5). Of note, our analysis showed that those identifying as Black, Hispanic, and Indigenous/First Nations were more likely to experience lower affluence and higher disadvantage scores (Supplemental Fig. 3 and Supplemental Table 6). Those identifying as two or more races/ethnicities have a slightly elevated incidence of high disadvantage, with a similar affluence score. Higher affluence scores and lower disadvantage scores were found for those identifying as Asian, South Asian, and Middle Eastern as compared to White.

To account for the impact of socioeconomic status in driving the disparities in symptom severity between racial and ethnic groups, after testing for multicollinearity we included affluence scores as a confounding variable in our linear model, and present this analysis separately in Fig. 2 (and Table 3). For the races and ethnicities that had lower socioeconomic status (Black, Hispanic, Indigenous, and two or more ethnicities), most symptoms had lowered odds ratios when accounting the affluence score. Overall, these shifts in odds ratios were small, with the largest change for Black and Indigenous/First Nations women being fatigue/low energy with decreases of 0.03 (OR=1.13, 97.5% CI[1.02,1.27]) and 0.08, (OR=1.57, 97.5% CI[1.15,2.22]) respectively. Unusual period was decreased the most for Hispanic women, by 0.03 (OR=1.44, 97.5% CI[1.25,1.66]), and those identifying as two or more ethnicities, by 0.01 (OR=1.32, 97.5% CI[1.10,1.60]). Within these groups there are some symptoms that have increased odds ratios; weight changes (OR=1.29, 97.5% CI[1.16,1.43]) for Hispanic women and mood swings (OR=1.21, 97.5% CI[1.09,1.35]), heart discomfort (OR=1.20, 97.5% CI[1.06,1.36]) and joint/muscular discomfort (OR=1.23, 97.5% CI[1.10,1.38]) for those identifying as two or more ethnicities. The odds ratios for weight changes increased for women identifying as two or more ethnicities, but it remains insignificant. It is worth noting the increases are small, with the largest being an increase of 0.003 for joint/muscular discomfort in those identifying as two or more ethnicities. The statistical significance of odds ratios was minimally impacted, with only hot flashes and skin/hair changes for Indigenous/First Nations women becoming non-significant. Interestingly, adjusting for affluence score increases the odds ratios for all symptoms among those with higher socioeconomic status: Asian, Middle Eastern, and South Asian. For those identifying as Asian, most symptoms remained non-significant with the exception of heart discomfort (OR=1.47, 97.5% CI[1.13,1.90]) remaining significant (p=0.004) and skin/hair changes (OR=1.26, 97.5% CI[1.02,1.57]) becoming significant (p=0.036). For those identifying as Middle Eastern, brain fog (OR=1.88, 97.5% CI[1.27,2.90]), vaginal dryness (OR=1.66, 97.5% CI[1.12,2.53]), skin/hair changes (OR=1.75, 97.5% CI[1.12,2.85]), fatigue/low energy (OR=2.14, 97.5% CI[1.26,3.99]), and weight changes (OR=2.22, 97.5% CI[1.24,4.38]) all remain statistically significant, but no additional symptoms became significant. For South Asian women, weight change (OR=3.02, 97.5% CI[1.56,6.78]) is still statistically significant and all other symptoms remain insignificant. Similar results were observed using disadvantage score in the model in place of affluence score (Supplemental Table 7).

**FIG. 2.**
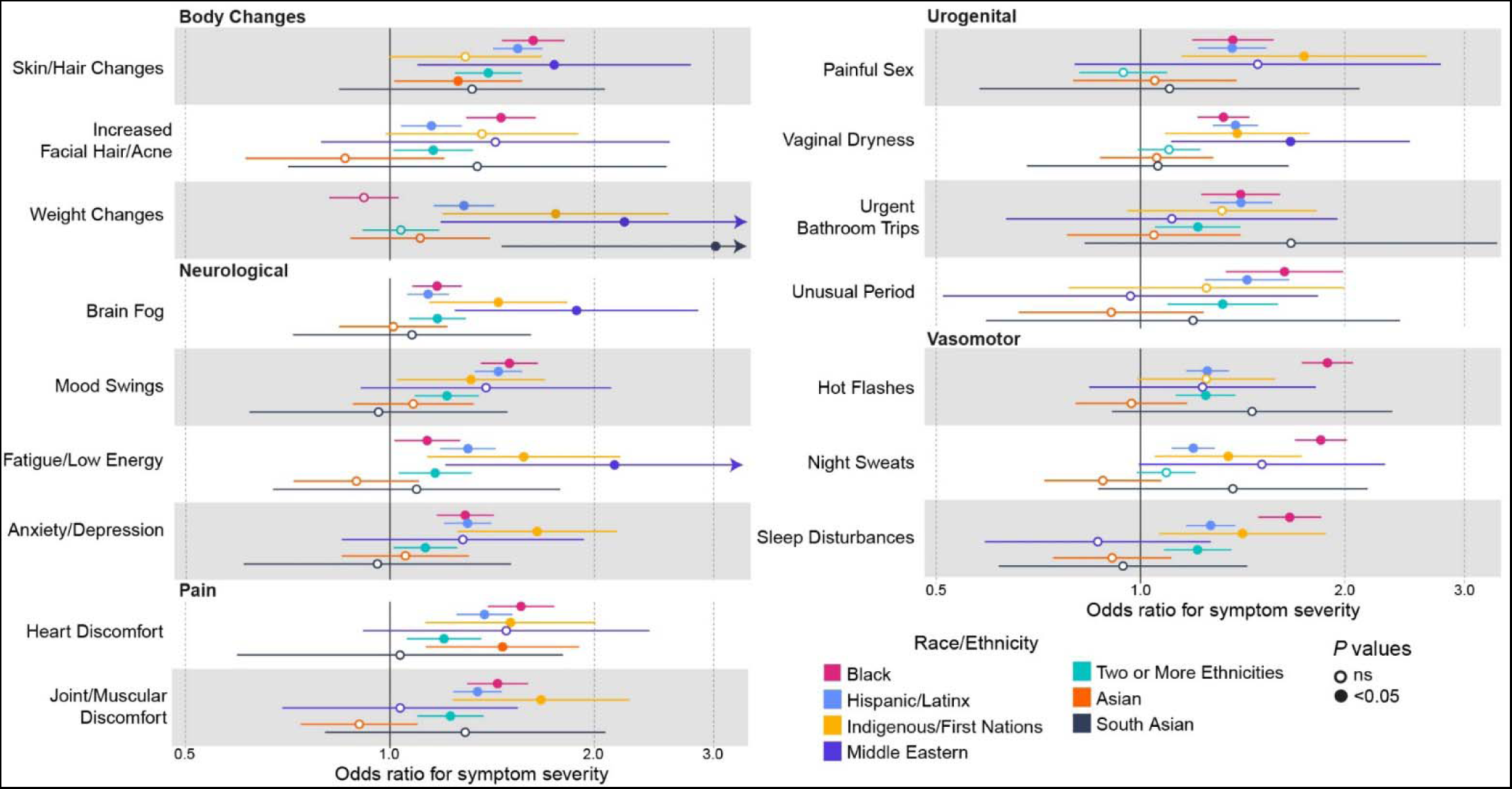
Adjusted Odds Ratios for Symptom Severity by Race/Ethnicity Including Affluence. Adjusted for Affluence score, age, BMI, smoking, and oophorectomy status. For unusual periods adjusted for all hysterectomy. Reference = White. Symptom Scores: 1-2 = Not Severe, 3-4 = Severe. Arrows indicate 95% CI extends beyond range.

**TABLE 3.** Adjusted Odds Ratios for Symptom Severity by Race and Ethnicity including Affluence Score^a^.

| Symptoms | Odds Ratio<br>(97.5% CI)<br>P value |  |  |  |  |  |  |
| --- | --- | --- | --- | --- | --- | --- | --- |
|  | Asian | Black | Hispanic/<br>Latinx | Indigenous/<br>First Nations | Middle<br>Eastern | South Asian | Two or More<br>Ethnicities |
| Hot Flashes | 0.97<br>(0.80-1.17)<br>P=.75 | **1.89**<br>**(1.73-2.06)**<br>**P<.001** | **1.25**<br>**(1.17-1.35)**<br>**P<.001** | 1.25<br>(0.99-1.59)<br>P=.06 | 1.23<br>(0.85-1.83)<br>P=.28 | 1.46<br>(0.92-2.39)<br>P=.12 | **1.25**<br>**(1.13-1.38)**<br>**P<.001** |
| Night Sweats | 0.88<br>(0.72-1.08)<br>P=.21 | **1.84**<br>**(1.69-2.01)**<br>**P<.001** | **1.20**<br>**(1.11-1.29)**<br>**P<.001** | **1.35**<br>**(1.05-1.74)**<br>**P=.02** | **1.51**<br>**(1.01-2.33)**<br>**P=.05** | 1.37<br>(0.88-2.20)<br>P=.18 | 1.09<br>(0.99-1.21)<br>P=.08 |
| Sleep Disturbances | 0.91<br>(0.75-1.11)<br>P=.35 | **1.66**<br>**(1.49-1.85)**<br>**P<.001** | **1.27**<br>**(1.17-1.38)**<br>**P<.001** | **1.41**<br>**(1.08-1.89)**<br>**P=.02** | 0.87<br>(0.60-1.29)<br>P=.46 | 0.94<br>(0.91-1.46)<br>P=.78 | **1.21**<br>**(1.08-1.36)**<br>**P=.001** |
| Brain Fog | 1.01<br>(0.84-1.22)<br>P=.90 | **1.17**<br>**(1.08-1.28)**<br>**P<.001** | **1.14**<br>**(1.06-1.22)**<br>**P<.001** | **1.44**<br>**(1.15-1.84)**<br>**P=.002** | **1.88**<br>**(1.27-2.90)**<br>**P=.003** | 1.08<br>(0.73-1.63)<br>P=.72 | **1.17**<br>**(1.07-1.29)**<br>**P=.001** |
| Painful Sex | 1.05<br>(0.80-1.39)<br>P=.73 | **1.37**<br>**(1.19-1.57)**<br>**P<.001** | **1.36**<br>**(1.22-1.53)**<br>**P<.001** | **1.74**<br>**(1.17-2.70)**<br>**P=.009** | 1.49<br>(0.83-2.91)<br>P=.21 | 1.10<br>(0.60-2.20)<br>P=.76 | 0.94<br>(0.81-1.10)<br>P=.44 |
| Vaginal Dryness | 1.06<br>(0.87-1.28)<br>P=.50 | **1.32**<br>**(1.21-1.45)**<br>**P<.001** | **1.38**<br>**(1.28-1.49)**<br>**P<.001** | **1.39**<br>**(1.09-1.78)**<br>**P=.009** | **1.66**<br>**(1.12-2.53)**<br>**P=.01** | 1.06<br>(0.69-1.67)<br>P=.79 | 1.10<br>(0.99-1.23)<br>P=.08 |
| Urgent Bathroom Trips | 1.05<br>(0.78-1.41)<br>P=.76 | **1.40**<br>**(1.23-1.61)**<br>**P<.001** | **1.41**<br>**(1.27-1.56)**<br>**P<.001** | 1.32<br>(0.96-1.83)<br>P=.09 | 1.11<br>(0.64-1.99)<br>P=.71 | 1.67<br>(0.85-3.50)<br>P=.15 | **1.21**<br>**(1.05-1.41)**<br>**P=.008** |
| Mood Swings | 1.08<br>(0.88-1.33)<br>P=.45 | **1.50**<br>**(1.36-1.65)**<br>**P<.001** | **1.44**<br>**(1.33-1.57)**<br>**P<.001** | **1.32**<br>**(1.03-1.70)**<br>**P=.03** | 1.39<br>(0.92-2.15)<br>P=.13 | 0.96<br>(0.63-1.51)<br>P=.86 | **1.44**<br>**(1.09-1.35)**<br>**P<.001** |
| Unusual Period | 0.91<br>(0.67-1.25)<br>P=.54 | **1.63**<br>**(1.34-2.00)**<br>**P<.001** | **1.44**<br>**(1.25-1.66)**<br>**P<.001** | 1.25<br>(0.80-2.03)<br>P=.34 | 0.97<br>(0.52-1.88)<br>P=.92 | 1.20<br>(0.61-2.52)<br>P=.62 | **1.32**<br>**(1.10-1.60)**<br>**P=.004** |
| Skin/Hair Changes | 1.26<br>(1.02-1.57)<br>P=.04 | **1.63**<br>**(1.46-1.81)**<br>**P<.001** | **1.54**<br>**(1.42-1.68)**<br>**P<.001** | **1.29**<br>**(1.00-1.68)**<br>**P=.05** | **1.75**<br>**(1.12-2.85)**<br>**P=.02** | 1.32<br>(0.86-2.11)<br>P=.23 | **1.40**<br>**(1.25-1.56)**<br>**P<.001** |
| Increased Facial Hair/Acne | 0.86<br>(0.61-1.21)<br>P=.38 | **1.46**<br>**(1.30-1.64)**<br>**P<.001** | **1.15**<br>**(1.04-1.28)**<br>**P=.007** | 1.37<br>(0.99-1.91)<br>P=.06 | 1.43<br>(0.81-2.65)<br>P=.24 | 1.35<br>(0.72-2.63)<br>P=.37 | **1.16**<br>**(1.01-1.33)**<br>**P=.03** |
| Fatigue/Low Energy | 0.89<br>(0.72-1.11)<br>P=.29 | **1.13**<br>**(1.02-1.27)**<br>**P=.03** | **1.30**<br>**(1.19-1.43)**<br>**P<.001** | **1.57**<br>**(1.15-2.22)**<br>**P=.007** | **2.14**<br>**(1.26-3.99)**<br>**P=.009** | 1.09<br>(0.69-1.83)<br>P=.72 | **1.17**<br>**(1.03-1.32)**<br>**P=.01** |
| Weight Changes | 1.11<br>(0.88-1.41)<br>P=.39 | 0.92<br>(0.82-1.03)<br>P=.14 | **1.29**<br>**(1.16-1.43)**<br>**P<.001** | **1.75**<br>**(1.22-2.63)**<br>**P=.004** | **2.22**<br>**(1.24-4.38)**<br>**P=.01** | **3.02**<br>**(1.56-6.78)**<br>**P=.003** | 1.04<br>(0.91-1.18)<br>P=.58 |
| Anxiety/Depression | 1.05<br>(0.85-1.31)<br>P=.63 | **1.29**<br>**(1.17-1.42)**<br>**P<.001** | **1.30**<br>**(1.20-1.41)**<br>**P<.001** | **1.65**<br>**(1.27-2.18)**<br>**P<.001** | 1.28<br>(0.86-1.96)<br>P=.24 | 0.96<br>(0.62-1.53)<br>P=.86 | **1.13**<br>**(1.01-1.26)**<br>**P=.03** |
| Heart Discomfort | **1.47**<br>**(1.13-1.90)**<br>**P=.004** | **1.56**<br>**(1.39-1.75)**<br>**P<.001** | **1.38**<br>**(1.25-1.52)**<br>**P<.001** | **1.51**<br>**(1.13-2.02)**<br>**P=.005** | 1.48<br>(0.92-2.43)<br>P=.11 | 1.04<br>(0.59-1.80)<br>P=.90 | **1.20**<br>**(1.06-1.36)**<br>**P=.004** |
| Joint/Muscular Discomfort | 0.90<br>(0.74-1.10)<br>P=.30 | **1.44**<br>**(1.30-1.60)**<br>**P<.001** | **1.35**<br>**(1.24-1.46)**<br>**P<.001** | **1.67**<br>**(1.25-2.28)**<br>**P=.001** | 1.04<br>(0.70-1.56)<br>P=.86 | 1.29<br>(0.82-2.12)<br>P=.29 | **1.23**<br>**(1.10-1.38)**<br>**P<.001** |

<sup>a</sup> Severity: 1-2 = Not Severe, 3-4 = Severe. Adjusted for Age, BMI, Smoking, Bilateral Oophorectomy, and Affluence Score. (For Unusual Period adjusted for Hysterectomy instead of bilateral oophorectomy). Significant (p<.05) values bolded.

## DISCUSSION

Our study is to our knowledge the largest and most comprehensive to date to demonstrate that the relationship between race and ethnicity and the severity of menopause symptoms is not solely explained by differences in socioeconomic status but is itself an independent factor. After controlling for several proxies of socioeconomic status, self-identified Black women, Hispanic, Indigenous or multi-racial or multi-ethnic groups have a higher incidence of worse menopause symptom severity. Our findings add to a growing body of evidence highlighting the strong connection between race and ethnicity, along with a lower socioeconomic status, and the severity of menopause symptoms^5^.

Several studies have shown there are racial and ethnic differences in the prevalence and severity of symptoms associated with the menopausal transition, with Black and Hispanic women reporting worse symptoms^5^. Prior literature has suggested women from lower socioeconomic backgrounds are more likely to experience worse menopausal symptoms than those from higher socioeconomic backgrounds^6^. To explore this further, our study aimed to determine whether adjusting for SES would eliminate racial and ethnic disparities in menopausal symptomatology. Even after controlling for proxies of SES, Black and Hispanic women still reported worse menopausal symptoms than non-Hispanic White women. This suggests factors outside of SES may be contributing to disparities in symptomatology.

Limited data exists on the menopause experiences of Indigenous women, but a systematic review revealed substantial variability in knowledge and symptom experience^14^. Indigenous women often experience an earlier onset of menopause attributed to factors such as poor nutrition and a harsher lifestyle. Our study suggests Indigenous women experience more severe menopause symptoms than White women, mirroring the symptom severity of Black and Hispanic women. Given limited access to care and increased discrimination, these findings highlight the need for further research and healthcare interventions to address the disparities in menopause symptom severity experienced by Indigenous women.

The menopause experiences of South Asian women living in the United States have received limited attention in the literature. A study conducted in Australia, which has a large South Asian population, found that women of Indian descent had a lower prevalence of classical menopausal symptoms compared to Caucasians, but reported higher levels of physical and psychological symptoms than the typical vasomotor symptoms^15^. Our study sheds new light on the symptom severity of South Asian women living in the United States, suggesting they have a similar level of symptomatology to White women, except for weight changes. South Asian women rated weight changes three times more severe than White women. The reason for this difference remains unclear. A plausible hypothesis is an increased rate of metabolic dysfunction in these patients, as seen in those with polycystic ovary syndrome^16^. Further research into this relationship is necessary to gain a deeper understanding of the menopause experiences of South Asian women.

The severity of menopausal symptoms varies among women and can be influenced by genetics, lifestyle, and health factors^17^. Cultural, social, and economic factors affecting access to healthcare and ability to manage symptoms can also contribute to disparities. For example, a study by Finley et al. suggested that women of moderate to high income were three times more likely than those of low income to use hormone replacement therapy^18^. It is also well documented in other areas of healthcare that Black and Hispanic women face significant barriers to accessing care and are more likely to discontinue treatment, barriers that can’t necessarily be controlled for with proxies for SES^19^. Differences in cultural attitudes, beliefs, and practices regarding menopause and aging may also contribute to disparities in symptom experience among women from diverse communities. Hormonal profiles may also differ among women of different ethnicities^20^.

This study highlights that while there is a correlation between race and ethnicity and the severity of menopausal symptoms, further research is needed to better understand the complexities of this issue, including cultural, lifestyle, genetic, or other factors. The authors caution against attributing differences solely to race and ethnicity, and highlights the need for a more nuanced understanding of this important health issue.

This is to our knowledge the largest study to date investigating menopause symptom severity by race or ethnicity when controlling for income and includes a robust sample size to examine symptomatology among different groups. Other strengths include controlling for important covariates, such as oophorectomy and hysterectomy status, which are contributors to menopausal symptoms, and incorporating a proxy for SES into the model as a covariate.

Limitations include the fact that all participants were English-speaking patients who sought care through the Evernow platform, so the results may not represent all individuals across different racial and ethnic groups nor should be taken to represent prevalence rates of symptomatology. Patients must had had at least one MRS symptom to be included in this study, which may bias the findings and generalizability of the findings. Patients self-reported their history and symptoms which can introduce bias.

Although we report it, we did not examine symptom prevalence in depth as the focus of this study was on symptom severity. Furthermore, the use of ZIP code data to control for SES may not accurately reflect an individual’s true SES and other measures of SES that were not accounted for could also be determinants of symptom severity, such as structural racism or geographic issues. Finally, it is important to note that this study only suggests associations and does not allow for the evaluation of causal relationships, necessitating further research to fully understand the findings.

## CONCLUSIONS

This study suggests that the severity of menopause symptoms differs by race and ethnicity regardless of SES and highlights the importance of considering race and ethnicity as independent factors in menopause experience. Importantly, certain ethnic groups may be at increased risk for adverse cardiovascular events due to frequent and persistent VMS and may require risk reduction strategies^21^. The study emphasizes the need to address social, cultural, and economic factors that contribute to disparities in menopausal symptoms and improve access to healthcare for all women with symptoms during this life stage.

## Summary Sentence

The relationship between race and ethnicity and menopause is not solely explained by differences in socioeconomic status, but is itself an independent factor.

## Data Availability

All data produced in the present study are available upon reasonable request to the authors

## Supplemental Digital Content

Supplemental Figures and Tables.docx

Supplemental Methods.docx

