## Supplemental Figures and Tables for "The Impact of Race, Ethnicity, and Socioeconomic Status on the Severity of Menopause Symptoms: A Study of 68,864 Women"

| **Supplemental Table 1. Percentages of participants within each race and ethnicity category experiencing severe symptoms.** | | | | | | | | |
| --- | --- | --- | --- | --- | --- | --- | --- | --- |
|  | **No.**  **(% Present)** | | | | | | | |
| **Symptoms** | **White** | **Asian** | **Black** | **Hispanic /Latinx** | **Indigenous/ First Nations** | **Middle Eastern** | **South Asian** | **Two or More Ethnicities** |
| Hot Flashes | 23335/34320 (68.0) | 321/485 (64.3) | 2910/3604 (80.7) | 3028/4183 (72.4) | 275/373 (73.7) | 88/126 (69.8) | 62/86  (72) | 1439/1993 (72.2) |
| Night Sweats | 23344/34260 (68.1) | 273/430 (63.5) | 2750/3437 (80.0) | 2830/3964 (71.4) | 252/336 (75.0) | 88/118 (74.6) | 66/92  (72) | 1363/1960 (69.5) |
| Sleep Disturbances | 34382/41909 (82.0) | 466/591 (78.8) | 3286/3696 (88.9) | 4193/4918 (85.3) | 385/441 (87.3) | 121/155 (78.1) | 101/129 (78.3) | 2023/2393 (84.5) |
| Brain Fog | 29803/41384 (72.0) | 390/556 (70.1) | 2545/3333 (76.4) | 3439/4595 (74.9) | 356/445 (80.0) | 121/149 (81.2) | 80/114 (70.2) | 1781/2366 (75.3) |
| Painful Sex | 15781/19673 (80.2) | 256/322 (79.5) | 1470/1737 (84.6) | 2013/2407 (83.6) | 185/211 (87.7) | 67/79  (85) | 45/57  (79) | 870/1113 (78.2) |
| Vaginal Dryness | 19924/29960 (66.5) | 314/480 (65.4) | 2073/2849 (72.8) | 2727/3766 (72.4) | 253/343 (73.8) | 94/126 (74.6) | 56/87  (64) | 1118/1661 (67.3) |
| Urgent Bathroom Trips | 10200/15337 (66.5) | 126/198 (63.6) | 975/1289 (75.6) | 1488/2020 (73.7) | 146/198 (73.7) | 36/55  (66) | 29/40  (73) | 674/954 (70.6) |
| Mood Swings | 22057/31459 (70.1) | 308/442 (69.7) | 2118/2689 (78.8) | 2934/3788 (77.5) | 272/353 (77.1) | 84/113 (74.3) | 59/90  (66) | 1343/1809 (74.2) |
| Unusual Period | 6632/9375 (70.7) | 123/183 (67.2) | 551/682 (80.8) | 959/1230 (78.0) | 75/99  (76) | 31/45  (69) | 28/39  (72) | 502/655 (76.6) |
| Skin/Hair  Changes | 23922/32119  (74.5) | 365/474 (77.0) | 2250/2698 (83.4) | 3364/4117 (81.7) | 293/366 (80.1) | 102/124 (82.3) | 82/107 (76.6) | 1613/2016 (80.0) |
| Increased Facial Hair/Acne | 12033/18452 (65.2) | 84/143 (58.7) | 1196/1602 (74.7) | 1269/1859 (68.3) | 139/189 (73.5) | 37/53  (70) | 29/43  (67) | 725/1054 (68.8) |
| Fatigue/Low Energy | 37207/43571 (85.4) | 459/569 (80.7) | 3078/3465 (88.8) | 4425/4973 (89.0) | 433/473 (91.5) | 130/143 (90.9) | 95/115 (82.6) | 2174/2482 (87.6) |
| Weight Changes | 36843/42440 (86.8) | 479/566 (84.6) | 3064/3414 (89.7) | 4406/4867 (90.5) | 408/437 (93.4) | 142/153 (92.8) | 115/123 (93.5) | 2105/2393 (88.0) |
| Anxiety/  Depression | 23782/33024 (72.0) | 291/410 (71.0) | 2040/2619 (77.9) | 2939/3810 (77.1) | 299/364 (82.1) | 90/121 (74.4) | 58/86  (67) | 1395/1871 (74.6) |
| Heart Discomfort | 8327/16838 (49.5) | 134/237 (56.5) | 878/1420 (61.8) | 1136/1973 (57.6) | 121/198 (61.1) | 38/67  (57) | 24/51  (47) | 560/1039 (53.9) |
| Joint/Muscular Discomfort | 26138/34279 (76.3) | 339/485 (69.9) | 2511/2984 (84.1) | 3425/4211 (81.3) | 313/364 (86.0) | 92/126 (73.0) | 71/94  (76) | 1635/2050 (79.8) |

| **Supplemental Table 2. Percentages of participants within each race and ethnicity category experiencing symptom presence** | | | | | | | | |
| --- | --- | --- | --- | --- | --- | --- | --- | --- |
|  | **No.**  **(% Present)** | | | | | | | |
| **Symptoms** | **White** | **Asian** | **Black** | **Hispanic/**  **Latinx** | **Indigenous/**  **First**  **Nations** | **Middle**  **Eastern** | **South**  **Asian** | **Two or**  **More**  **Ethnicities** |
| Hot Flashes | 34320/52514 (65.4) | 485/790 (61.4) | 3604/4579 (78.7) | 4183/6116 (68.4) | 373/531 (70.2) | 126/206 (61.2) | 86/161 (53.4) | 1993/2970 (67.1) |
| Night Sweats | 34260/52512 (65.2) | 430/790 (54.4) | 3437/4579 (75.1) | 3964/6116 (64.8) | 336/531 (63.3) | 118/206 (57.3) | 92/161 (57.1) | 1960/2970 (66.0) |
| Sleep Disturbances | 41909/52514 (79.8) | 591/790 (74.8) | 3696/4579 (80.7) | 4918/6116 (80.4) | 441/531 (83.1) | 115/206 (75.2) | 129/161 (80.1) | 2393/2970 (80.6) |
| Brain Fog | 41384/52514 (78.8) | 556/790 (70.4) | 3333/4579 (72.8) | 4594/6116 (75.1) | 445/531 (83.8) | 149/206 (72.3) | 114/161 (70.8) | 2366/2970 (79.7) |
| Painful Sex | 19673/52514 (37.5) | 322/790 (40.8) | 1737/4579 (37.9) | 2407/6116 (39.4) | 211/531 (39.7) | 79/206 (38.3) | 57/161 (35.4) | 1113/2970 (37.5) |
| Vaginal Dryness | 29960/52514 (57.0) | 480/790 (60.8) | 2849/4579 (62.2) | 3766/6116 (61.6) | 343/531 (64.6) | 126/206 (61.2) | 87/161 (54.0) | 1661/2970 (55.9) |
| Urgent  Bathroom Trips | 15337/52514 (29.2) | 198/790 (25.1) | 1289/4579 (28.2) | 2020/6116 (33.0) | 198/531 (37.3) | 55/206 (26.7) | 40/161 (24.8) | 954/2970 (32.1) |
| Mood Swings | 31459/52514 (59.9) | 442/790 (55.9) | 2589/4579 (58.7) | 3788/6116 (61.9) | 353/531 (66.5) | 113/206 (54.9) | 90/161 (55.9) | 1809/2970 (60.9) |
| Unusual Period | 9375/52514 (17.9) | 183/790 (23.2) | 682/4579 (14.9) | 1230/6116 (20.1) | 99/531 (18.6) | 45/206 (21.8) | 39/161 (24.2) | 655/2970 (22.1) |
| Skin/Hair  Changes | 32119/52514 (61.2) | 474/790 (60.0) | 2698/4579 (58.9) | 4117/6116 (67.3) | 366/531 (68.9) | 124/206 (60.2) | 107/161 (66.5) | 2016/2970 (67.9) |
| Increased  Facial  Hair/Acne | 18452/52514 (35.2) | 143/790 (18.2) | 1602/4579 (35.0) | 1859/6116 (30.4) | 189/531 (35.6) | 53/206 (25.7) | 43/161 (26.7) | 1054/2970 (35.5) |
| Fatigue/Low Energy | 43571/52514 (83.0) | 569/790 (72.0) | 3465/4579 (75.7) | 4973/6116 (81.3) | 473/531 (89.0) | 143/206 (69.4) | 115/161 (71.4) | 2482/2970 (83.6) |
| Weight  Changes | 42440/52514 (80.8) | 566/790 (71.6) | 3414/4579 (74.6) | 4867/6116 (79.6) | 437/531 (82.3) | 153/206 (74.3) | 123/161 (76.4) | 2393/2970 (80.6) |
| Anxiety/  Depression | 33024/52514 (62.9) | 410/790 (51.9) | 2619/4579 (57.2) | 3810/6116 (62.3) | 364/531 (68.5) | 121/206 (58.7) | 86/161 (53.4) | 1871/2970 (63.0) |
| Heart  Discomfort | 16838/52514 (32.1) | 237/790 (30.0) | 1420/4579 (32.3) | 1973/6116 (32.3) | 198/531 (37.3) | 67/206 (32.5) | 51/161 (31.7) | 1039/2970 (35.0) |
| Joint/Muscular Discomfort | 34279/52514 (65.3) | 485/790 (61.4) | 2984/4579 (65.2) | 4211/6116 (68.9) | 364/531 (68.5) | 126/206 (61.2) | 94/161 (58.4) | 2050/2970 (69.0) |

| **Supplemental Table 3. Mean Affluence and Disadvantage Scores for US Census Data and Survey Participants** | | | |
| --- | --- | --- | --- |
|  | **Mean** | **SEM** | **n** |
| NaNDA Affluence Score | 0.326 | 0.001 | 320990935 |
| NaNDA Disadvantage Score | 0.099 | 0.099 | 320990935 |
| Evernow Affluence Score | 0.398 | 0.001 | 67790 |
| Evernow Disadvantage Score | 0.094 | 0.000 | 67791 |

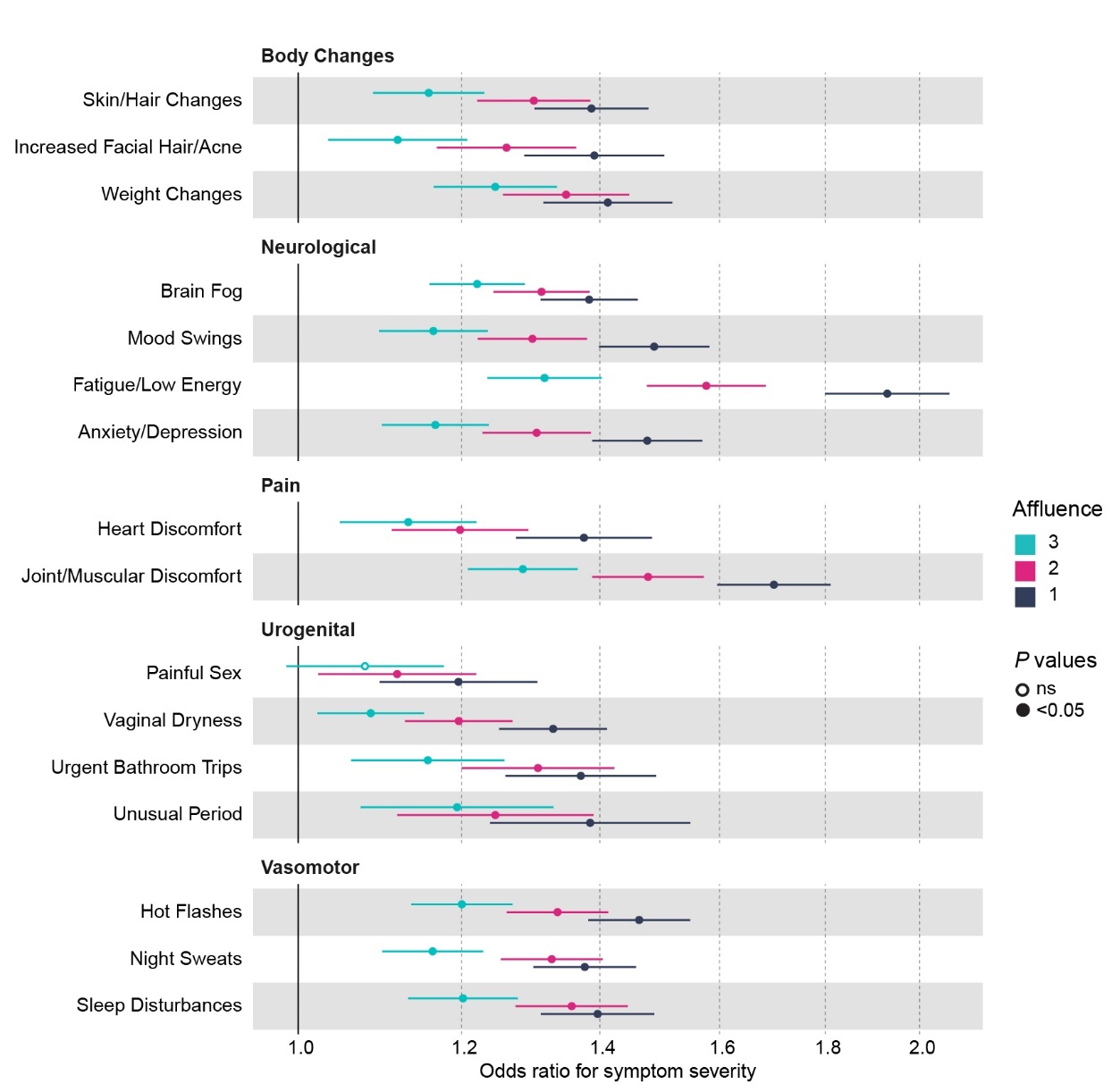

**Supplemental Fig. 1. Odds Ratios for Symptom Severity by Affluence Score**

Unadjusted odds ratios. Affluence Score: 1 = Low Affluence, 4 = High Affluence. Reference = 4. Symptom Scores: 1-2 = Not Severe, 3-4 = Severe.

| **Supplemental Table 4. Unadjusted Odds Ratios for Symptom Severity (1-2 = Not Severe, 3-4 = Severe) by Affluence (1 = low affluence, 4 = high affluence). OR = odds ratio.** | | | | | | | | | | |
| --- | --- | --- | --- | --- | --- | --- | --- | --- | --- | --- |
|  | **Score 4** | **Score 1** | | | **Score 2** | | | **Score 3** | | |
| **Symptoms** | **% Severe** | **% Severe** | **OR**  **(97.5% CI)** | ***P* value** | **% Severe** | **OR**  **(97.5% CI)** | ***P* value** | **% Severe** | **OR**  **(97.5% CI)** | ***P* value** |
| Hot Flashes | 6951/10706 (64.9) | 8581/11750 (73.0) | 1.46  (1.38-1.55) | P<.001 | 8215/11538 (71.2) | 1.34  (1.26-1.41) | P<.001 | 7677/11132 (69.0) | 1.20  (1.13-1.27) | P<.001 |
| Night Sweats | 7076/10830 (65.3) | 8104/11227 (72.2) | 1.38  (1.30-1.46) | P<.001 | 8084/11316 (71.4) | 1.33  (1.25-1.40) | P<.001 | 7672/11175 (68.7) | 1.16  (1.10-1.23) | P<.001 |
| Sleep Disturbances | 10803/13521 (79.9) | 11352/13397 (84.7) | 1.40  (1.31-1.49) | P<.001 | 11476/13604 (84.4) | 1.36  (1.27-1.44) | P<.001 | 11284/13646 (82.7) | 1.20  (1.13-1.28) | P<.001 |
| Brain Fog | 8993/13090 (68.7) | 9906/13168 (75.2) | 1.38  (1.31-1.46) | P<.001 | 9840/13257 (74.2) | 1.31  (1.24-1.38) | P<.001 | 9734/13366 (72.8) | 1.22  (1.16-1.29) | P<.001 |
| Painful Sex | 4818/6071 (79.4) | 5527/6729 (82.1) | 1.20  (1.10-1.31) | P<.001 | 5192/6401  (81.1) | 1.12  (1.02-1.22) | 0.01 | 5125/6362 (80.6) | 1.08  (0.99-1.18) | 0.10 |
| Vaginal Dryness | 6115/9467 (64.6) | 7123/10061 (70.8) | 1.33  (1.25-1.41) | P<.001 | 6776/9881  (68.6) | 1.20  (1.13-1.27) | P<.001 | 6520/9816 (66.4) | 1.08  (1.02-1.15) | 0.008 |
| Urgent  Bathroom  Trips | 2838/4445 (63.8) | 3936/5562 (70.8) | 1.37  (1.26-1.49) | P<.001 | 3593/5150  (69.8) | 1.31  (1.20-1.42) | P<.001 | 3294/4908 (67.1) | 1.16  (1.06-1.26) | 0.001 |
| Mood Swings | 6344/9427 (67.3) | 8096/10741 (75.4) | 1.49  (1.40-1.58) | P<.001 | 7533/10352 (72.8) | 1.30  (1.22-1.38) | P<.001 | 7176/10175 (70.5) | 1.16  (1.09-1.24) | P<.001 |
| Unusual Period | 2327/3381 (68.8) | 2171/2881 (75.4) | 1.38  (1.24-1.55) | P<.001 | 2154/2937  (73.3) | 1.25  (1.12-1.39) | P<.001 | 2238/3087 (72.5) | 1.19  (1.07-1.33) | 0.001 |
| Skin/Hair Changes | 7515/10336 (72.7) | 8295/10540 (78.7) | 1.39  (1.30-1.48) | P<.001 | 8153/10506 (77.6) | 1.30  (1.22-1.39) | P<.001 | 7991/10584 (75.5) | 1.16  (1.09-1.23) | P<.001 |
| Increased  Facial  Hair/Acne | 3240/5198 (62.3) | 4301/6169 (69.7) | 1.39  (1.29-1.50) | P<.001 | 4142/6126  (67.6) | 1.26  (1.17-1.36) | P<.001 | 3809/5869 (64.9) | 1.12  (1.03-1.21) | 0.005 |
| Fatigue/Low Energy | 11008/13490 (81.6) | 12534/13999 (89.5) | 1.93  (1.80-2.07) | P<.001 | 12327/14090 (87.5) | 1.58  (1.48-1.68) | P<.001 | 12079/14148 (85.4) | 1.32  (1.24-1.40) | P<.001 |
| Weight Changes | 11699/13765 (85.0) | 11630/13084 (88.9) | 1.41  (1.31-1.52) | P<.001 | 11934/13497 (88.4) | 1.35  (1.26-1.45) | P<.001 | 12248/13984 (87.6) | 1.25  (1.16-1.33) | P<.001 |
| Anxiety/  Depression | 6973/10120 (68.9) | 8252/10775 (76.6) | 1.48  (1.39-1.57) | P<.001 | 7968/10724  (74.3) | 1.30  (1.23-1.39) | P<.001 | 7667/10636 (72.1) | 1.17  (1.10-1.24) | P<.001 |
| Heart Discomfort | 2429/5119 (47.5) | 3156/5697 (55.4) | 1.38  (1.28-1.48) | P<.001 | 2859/5502  (52.0) | 1.20  (1.11-1.29) | P<.001 | 2769/5481 (50.5) | 1.13  (1.05-1.22) | 0.002 |
| Joint/Muscular Discomfort | 7546/10484 (72.0) | 9431/11591 (81.4) | 1.70  (1.60-1.81) | P<.001 | 8974/11339 (79.1) | 1.48  (1.39-1.57) | P<.001 | 8537/11124 (76.7) | 1.28  (1.21-1.37) | P<.001 |

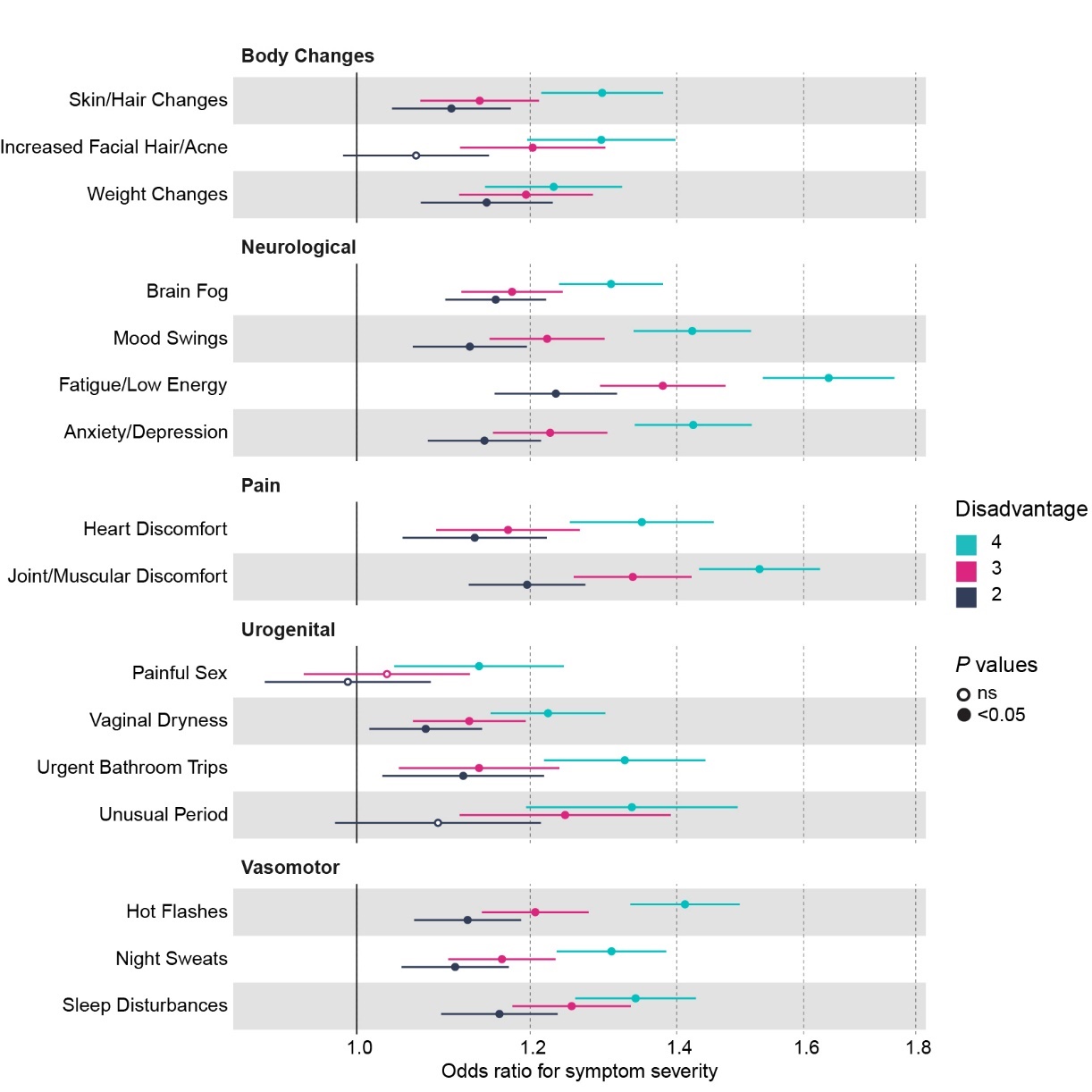

**Supplemental Fig. 2. Odds Ratios for Symptom Severity by Disadvantage Score**

Unadjusted odds ratios. Disadvantage Score: 1 = Low Disadvantage, 4 = High Disadvantage. Reference = 1. Symptom Scores: 1-2 = Not Severe, 3-4 = Severe.

| **Supplemental Table 5. Unadjusted Odds Ratios for Symptom Severity (1-2 = Not Severe, 3-4 = Severe) by Disadvantage (1 = low disadvantage, 4 = high disadvantage). OR = odds ratio.** | | | | | | | | | | |
| --- | --- | --- | --- | --- | --- | --- | --- | --- | --- | --- |
|  | **Score 1** | **Score 2** | | | **Score 3** | | | **Score 4** | | |
| **Symptoms** | **% Severe** | **% Severe** | **OR**  **(97.5% CI)** | ***P* value** | **% Severe** | **OR**  **(97.5% CI)** | ***P* value** | **% Severe** | **OR**  **(97.5% CI)** | ***P* value** |
| Hot Flashes | 7258/10976 (66.1) | 7742/11271 (68.7) | 1.12  (1.06-1.19) | P<.001 | 8068/11493 (70.2) | 1.21  (1.14-1.28) | P<.001 | 8355/11385 (73.4) | 1.41  (1.33-1.50) | P<.001 |
| Night Sweats | 7347/11026 (66.6) | 7715/11198 (68.9) | 1.11  (1.05-1.17) | P<.001 | 7925/11331 (69.9) | 1.16  (1.10-1.23) | P<.001 | 7948/10992 (72.3) | 1.31  (1.23-1.38) | P<.001 |
| Sleep  Disturbances | 11122/13815 (80.5) | 11320/13679 (82.8) | 1.16  (1.09-1.24) | P<.001 | 11364/13559 (83.8) | 1.25  (1.18-1.33) | P<.001 | 11108/13114 (84.7) | 1.34  (1.26-1.43) | P<.001 |
| Brain Fog | 9387/13433 (69.9) | 9791/13437 (72.9) | 1.16  (1.10-1.22) | P<.001 | 9712/13267 (73.2) | 1.18  (1.12-1.24) | P<.001 | 9583/12744 (75.2) | 1.31  (1.24-1.38) | P<.001 |
| Painful Sex | 5071/6319 (80.3) | 5104/6372 (80.1) | 0.99  0.91-1.08) | 0.83 | 5264/6519 (80.7) | 1.03  (0.95-1.13) | 0.48 | 5222/6352 (82.2) | 1.14  (1.04-1.24) | 0.005 |
| Vaginal Dryness | 6380/9742 (65.5) | 6611/9851 (67.1) | 1.08  (1.01-1.14) | 0.02 | 6810/9998 (68.1) | 1.13  (1.06-1.19) | P<.001 | 6732/9633 (69.9) | 1.22  (1.15-1.30) | P<.001 |
| Urgent  Bathroom Trips | 2979/4576 (65.1) | 3355/4963 (67.6) | 1.12  (1.03-1.22) | 0.01 | 3509/5163 (68.0) | 1.14  (1.05-1.24) | 0.003 | 3818/5362 (71.2) | 1.33  (1.22-1.44) | P<.001 |
| Mood Swings | 6707/9844 (68.1) | 7222/10222 (70.7) | 1.13  (1.06-1.20) | P<.001 | 7473/10335 (72.3) | 1.22  (1.15-1.30) | P<.001 | 7747/10294 (75.3) | 1.42  (1.34-1.51) | P<.001 |
| Unusual Period | 2252/3244 (69.4) | 2176/3056 (71.2) | 1.09  (0.98-1.21) | 0.12 | 2182/2954 (73.9) | 1.25  (1.11-1.39) | P<.001 | 2280/3032 (75.2) | 1.34  (1.20-1.49) | P<.001 |
| Skin/Hair Changes | 7780/10526 (73.9) | 7941/10478 (75.8) | 1.10  (1.04-1.18) | 0.002 | 8094/10604 (76.3) | 1.14  (1.07-1.21) | P<.001 | 8139/10358 (78.6) | 1.29  (1.21-1.38) | P<.001 |
| Increased Facial Hair/Acne | 3527/5561 (63.4) | 3767/5808 (64.9) | 1.06  (0.99-1.15) | 0.11 | 4148/6136 (67.6) | 1.20  (1.11-1.30) | P<.001 | 4050/5856 (69.2) | 1.29  (1.20-1.40) | P<.001 |
| Fatigue/Low Energy | 11600/14000 (82.9) | 12103/14134 (85.6) | 1.23  (1.16-1.31) | P<.001 | 12217/14049 (87.0) | 1.38  (1.29-1.47) | P<.001 | 12028/13543 (88.8) | 1.64  (1.53-1.76) | P<.001 |
| Weight Changes | 12090/14058 (86.0) | 12164/13891 (87.6) | 1.15  (1.07-1.23) | P<.001 | 11988/13621 (88.0) | 1.19  (1.11-1.28) | P<.001 | 11268/12759 (88.3) | 1.23  (1.14-1.32) | P<.001 |
| Anxiety/  Depression | 7253/10426 (69.6) | 7718/10670 (72.3) | 1.14  (1.08-1.21) | P<.001 | 7833/10629 (73.7) | 1.23  (1.15-1.30) | P<.001 | 8056/10530 (76.5) | 1.42  (1.34-1.51) | P<.001 |
| Heart Discomfort | 2523/5282 (47.8) | 2758/5423 (50.9) | 1.13  (1.05-1.22) | 0.001 | 2881/5569 (51.7) | 1.17  (1.09-1.26) | P<.001 | 3051/5524 (55.2) | 1.35  (1.25-1.46) | P<.001 |
| Joint/Muscular Discomfort | 7907/10776 (73.4) | 8553/11147 (76.7) | 1.20  (1.13-1.27) | P<.001 | 8976/11412 (78.7) | 1.34  (1.26-1.42) | P<.001 | 9052/11202 (80.8) | 1.53  (1.43-1.63) | P<.001 |

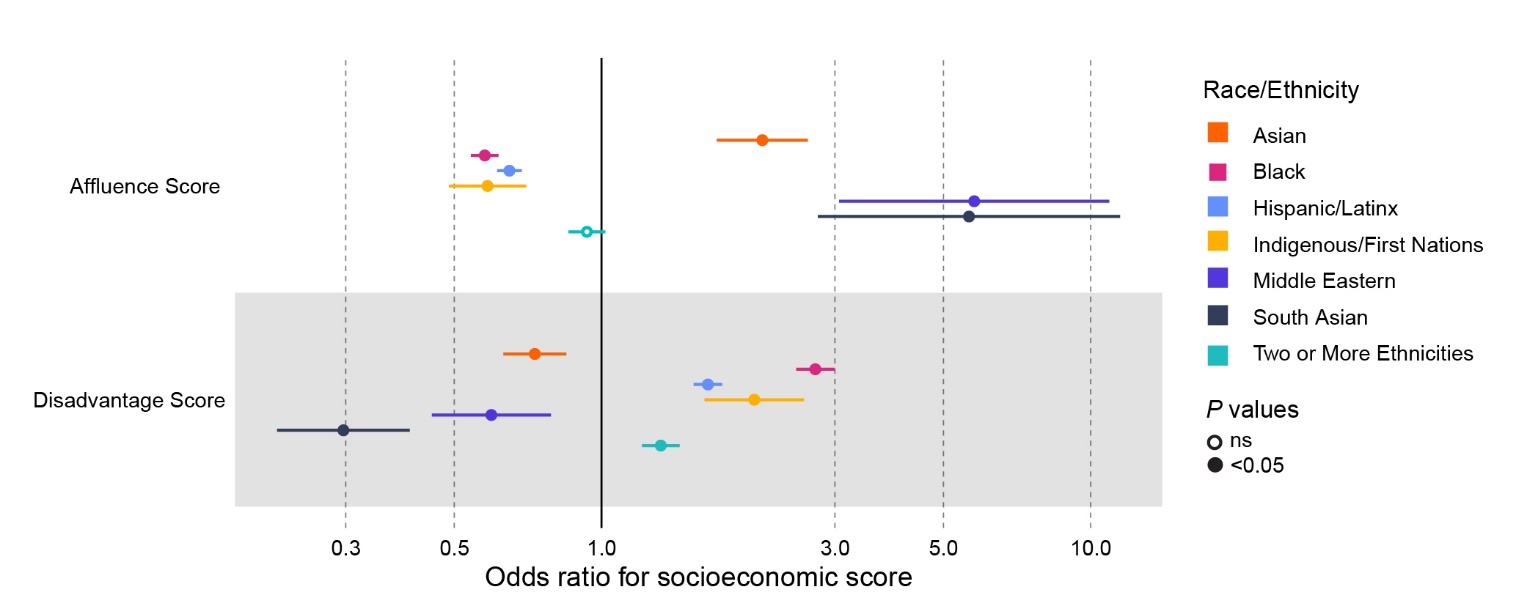

**Supplemental Figure 3. Odds ratios for Socioeconomic Scores by Race and Ethnicity.**

Unadjusted odds ratios. Reference = White.

| **Supplemental Table 6. Unadjusted Odds Ratios for Affluence and Disadvantage Scores by Race and Ethnicity. OR = odds ratio** | | | | | | | |
| --- | --- | --- | --- | --- | --- | --- | --- |
|  |  |  |  | **Odds Ratio**  **(97.5% CI)**  ***P* value** |  |  |  |
|  | **Asian** | **Black** | **Hispanic/**  **Latinx** | **Indigenous/ First Nations** | **Middle Eastern** | **South Asian** | **Two or More Ethnicities** |
| Affluence  Score | 2.1  (1.7-2.7)  P<.001 | 0.58  (0.54-0.62)  P<.001 | 0.64  (0.61 -0.69)  P<.001 | 0.58  (0.49-0.70)  P<.001 | 5.78  (3.23-11.68)  P<.001 | 5.64  (2.96-12.52)  P<.001 | 0.93  (0.86-1.02) |
| Disadvantage Score | 0.7  (0.6-0.8)  P<.001 | 2.74  (2.50-3.00)  P<.001 | 1.65  (1.54-1.76)  P<.001 | 2.05  (1.63-2.61)  P<.001 | 0.60  (0.45-0.79)  P<.001 | 0.30  (0.22-0.40)  P<.001 | 1.32  (1.21-1.44) |

| **Supplemental Table 7. Adjusted Odds Ratios for Symptom Severity (1-2 = Not Severe, 3-4 = Severe) by Race and Ethnicity including Disadvantage Score.** Adjusted for Age, BMI, Smoking, Bilateral Oophorectomy, and Disadvantage Score. (For Unusual Period adjusted for Hysterectomy instead of bilateral oophorectomy). OR = odds ratio. | | | | | | | |
| --- | --- | --- | --- | --- | --- | --- | --- |
|  | **Odds Ratio**  **(97.5% CI)**  ***P* value** | | | | | | |
| **Symptoms** | **Asian** | **Black** | **Hispanic/**  **Latinx** | **Indigenous/**  **First Nations** | **Middle**  **Eastern** | **South Asian** | **Two or More Ethnicities** |
| Hot Flashes | 0.95  (0.79-1.15)  *P*=.57 | 1.83  (1.68-2.00)  *P*<.001 | 1.24  (1.15-1.33)  *P*<.001 | 1.26  (1.00-1.59)  *P*=.05 | 1.16  (0.80-1.72)  *P*=.45 | 1.39  (0.88-2.27)  *P*=.18 | 1.23  (1.12-1.37)  *P*<.001 |
| Night Sweats | 0.86  (0.71-1.05)  *P*=.14 | 1.81  (1.66-1.98)  *P*<.001 | 1.19  (1.11-1.28)  *P*<.001 | 1.36  (1.06-1.75)  *P*=.02 | 1.43  (0.95-2.20)  *P*=.09 | 1.30  (0.83-2.09)  *P*=.26 | 1.08  (0.98-1.20)  *P*=.12 |
| Sleep Disturbances | 0.89  (0.73-1.09)  *P*=.27 | 1.63  (1.47-1.82)  *P*<.001 | 1.26  (1.16-1.37)  *P*<.001 | 1.42  (1.08-1.90)  *P*=.02 | 0.82  (0.57-1.22)  *P*=.31 | 0.91  (0.60-1.41)  *P*=.65 | 1.20  (1.07-1.35)  *P*=.002 |
| Brain Fog | 0.99  (0.83-1.19)  *P*=.92 | 1.15  (1.06-1.25)  *P*<.001 | 1.13  (1.05-1.21)  *P*=.001 | 1.45  (1.15-1.84)  *P*=.002 | 1.78  (1.20-2.74)  *P*=.006 | 1.03  (0.69-1.56)  *P*=.89 | 1.16  (1.06-1.28)  *P*=.002 |
| Painful Sex | 1.04  (0.79-1.38)  *P*=.79 | 1.35  (1.17-1.55)  *P*<.001 | 1.36  (1.21-1.53)  *P*<.001 | 1.75  (1.18-2.71)  *P*=.009 | 1.45  (0.81-2.83)  *P*=.24 | 1.07  (0.58-2.12)  *P*=.85 | 0.94  (0.81-1.09)  *P*=.42 |
| Vaginal Dryness | 1.04  (0.86-1.26)  *P*=.70 | 1.31  (1.20-1.43)  *P*<.001 | 1.38  (1.28-1.49)  *P*<.001 | 1.40  (1.10-1.79)  *P*=.008 | 1.59  (1.07-2.42)  *P*=.025 | 1.02  (0.66-1.61)  *P*=.92 | 1.09  (2.98-1.22)  *P*=.10 |
| Urgent  Bathroom Trips | 1.03  (0.77-1.39)  *P*=.85 | 1.38  (1.20-1.58)  *P*<.001 | 1.40  (1.26-1.55)  *P*<.001 | 1.32  (0.97-1.84)  *P*=.09 | 1.05  (0.60-1.87)  *P*=.87 | 1.59  (0.81-3.34)  *P*=.19 | 1.21  (1.04-1.39)  *P*=.01 |
| Mood Swings | 1.06  (0.86-1.30)  *P*=.59 | 1.46  (1.32-1.61)  *P*<.001 | 1.43  (1.32-1.55)  *P*<.001 | 1.31  (1.03-1.70)  *P*=.03 | 1.31  (0.87-2.03)  *P*=.21 | 0.92  (0.60-1.44)  *P*=.72 | 1.20  (1.08-1.34)  *P*=.001 |
| Unusual Period | 0.90  (0.66-1.23)  *P*=.49 | 1.60  (1.31-1.96)  *P*<.001 | 1.43  (1.24-1.66)  *P*<.001 | 1.25  (0.80-2.04)  *P*=.35 | 0.94  (0.51-1.82)  *P*=.84 | 1.16  (0.59-2.43)  *P*=.69 | 1.31  (1.09-1.59)  *P*=.004 |
| Skin/Hair  Changes | 1.24  (1.00-1.54)  *P*=.05 | 1.60  (1.44-1.79)  *P*<.001 | 1.54  (1.42-1.67)  *P*<.001 | 1.31  (1.01-1.70)  *P*=.04 | 1.65  (1.06-2.69)  *P*=.03 | 1.25  (0.81-2.00)  *P*=.33 | 1.39  (1.24-1.55)  *P*<.001 |
| Increased Facial Hair/Acne | 0.84  (0.60-1.18)  *P*=.31 | 1.43  (1.27-1.62)  *P*<.001 | 1.14  (1.03-1.27)  *P*=.01 | 1.37  (1.00-1.92)  *P*=.06 | 1.37  (0.78-2.55)  *P*=.29 | 1.29  (0.69-2.53)  *P*=.43 | 1.15  (1.01-1.32)  *P*=.04 |
| Fatigue/  Low Energy | 0.86  (0.70-1.07)  *P*=.17 | 1.10  (0.99-1.23)  *P*=.09 | 1.29  (1.18-1.42)  *P*<.001 | 1.59  (1.17-2.25)  *P*=.005 | 1.96  (1.15-3.65)  *P*=.02 | 1.03  (0.64-1.72)  *P*=.92 | 1.15  (1.02-1.30)  *P*=.03 |
| Weight Changes | 1.10  (0.87-1.40)  *P*=.43 | 0.93  (0.83-1.05)  *P*=.22 | 1.29  (1.17-1.44)  *P*<.001 | 1.77  (1.23-2.66)  *P*=.003 | 2.15  (1.21-4.26)  *P*=.02 | 2.96  (1.53-6.64)  *P*=.003 | 1.04  (0.91-1.19)  *P*=.55 |
| Anxiety/  Depression | 1.03  (0.83-1.28)  *P*=.80 | 1.25  (1.14-1.38)  *P*<.001 | 1.29  (1.19-1.39)  *P*<.001 | 1.65  (1.27-2.17)  *P*<.001 | 1.21  (0.81-1.85)  *P*=.36 | 0.92  (0.59-1.47)  *P*=.72 | 1.11  (1.00-1.24)  *P*=.05 |
| Heart Discomfort | 1.43  (1.11-1.86)  *P*=.007 | 1.53  (1.36-1.71)  *P*<.001 | 1.37  (1.24-1.50)  *P*<.001 | 1.50  (1.13-2.01)  *P*=.006 | 1.42  (0.88-2.33)  *P*=.16 | 1.01  (0.58-1.75)  *P*=.97 | 1.19  (1.05-1.35)  *P*=.007 |
| Joint/Muscular Discomfort | 0.88  (0.72-1.07)  *P*=.19 | 1.40  (1.26-1.56)  *P*<.001 | 1.34  (1.23-1.45)  *P*<.001 | 1.68  (1.26-2.29)  *P*=.001 | 0.97  (0.66-1.46)  *P*=.87 | 1.22  (0.77-2.00)  *P*=.41 | 1.21  (1.08-1.36)  *P*=.001 |
