## Supplemental Methods for "The Impact of Race, Ethnicity, and Socioeconomic Status on the Severity of Menopause Symptoms: A Study of 68,864 Women"

**Supplemental Methods: Survey Questions**

**Your Information**

What sex were you assigned at birth?

- Female
- Male

What is your gender identity? (select an option)

- Female
- Male
- Trans Female
- Trans Male
- Genderqueer/Non-conforming
- Different Identity

What is your legal date of birth?

Month/Day/Year

How tall are you?

Feet/Inches

What is your weight?

Weight(pounds)

Which of the following best describes you? (select all that apply)

- African
- African American
- Asian-American
- East Asian (including Chinese, Japanese, Korean, Mongolian, Tibetan, and Taiwanese)
- South Asian (including Bangladeshi, Bhutanese, Indian, Nepali, Pakistani, and Sri Lankan)
- Southeast Asian (including Burmese, Cambodian, Filipino, Hmong, Indonesian, Laotian, Mien, Singaporean, Thai, and Vietnamese)
- Hispanic/LatinX
- Indigenous American/First Nations (including Native American/American Indian, Alaskan Native, Pacific Islander and Native Hawaiian)
- Middle Eastern
- White
- Prefer not to say
- Other

**Your Symptoms**

What symptoms are you experiencing? (select all that apply)

- Hot flashes - Do you ever get waves of heat that come from nowhere and cause you to want to strip off your clothes? Do you feel the sudden need for an electric fan to keep cool?
- Night sweats - Do you ever wake up with the sheets / nightclothes drenched in sweat? Do you kick off the bedding in the middle of the night because you are sweaty?
- Vaginal dryness - Is your vagina always itchy / irritated? (and you don’t have a yeast infection)? Do you feel like it sometimes hurts if your vagina is touched: eg during pelvic exam / sexual activity?
- Problems Sleeping - Is it hard to fall asleep at night? Are you waking up in the middle of the night (maybe because of a night sweat)?
- Brain Fog - Do you ever feel like you make a list of things to do and then suddenly you can’t remember the last item? Is it harder for you to focus and finish tasks?
- Painful Sex - Do you feel like it is hard to stay lubricated during sexual activity? Do you feel like there is burning or pins and needles sensation with penetration? Do you feel like your vagina is now narrower than it used to be? Are you less interested in sex these days? Harder to climax?
- Urinary Problems - Do you feel like you always need to know where the bathroom is because you need to pee more frequently these days? Is it harder to hold your urine / pee? Do you lose urine when you cough / exercise? Do you have recurrent urinary tract infections?
- Mood Swings - Do you feel like your “fuse” is shorter these days? (Shorter temper). Are you more irritable and impatient? Feeling more aggressive?
- Period Irregularities - Period irregularities can include: light spotting, shorter cycles, skipped cycles, and bleeding between periods.
- Skin or Hair Changes - Is your hair thinner than before? Is your skin drier? Are you using a ton of night cream just to stay feeling normal?
- Facial Hair or Acne - Are you starting to get those random dark hairs that “just appear” on your face? That aggravating little zit?
- Fatigue - Are you always tired? Do you feel like you don’t have the energy to get your stuff / work done and just want to go to bed at home?
- Weight Changes - Do you feel like you are dieting / working out more these days but the scale doesn’t change?
- Anxiety or Depression - Are you feeling more nervous or panicky these days? Are you feeling more down or teary these days?
- Heart Discomfort - Do you have unusual awareness of your heart beat? Do you feel it skipping or racing?
- Muscle & Joint Pain - Do you feel stiffness, swelling, shooting pains, or even a burning sensation in your joints or muscles?

How severe are your symptoms? (Please rate your symptoms on a scale from 1=Mild to 4=Most Severe.)

**Health History**

Have you had a hysterectomy?

- Yes
- No

Why was the hysterectomy performed?

Were your ovaries removed?

- Yes
- No
- I’m not sure

How many ovaries were removed?

- One
- Both
- I don’t know

Have you ever had any other surgeries? For example, endometrial ablation or gallbladder removal

- Yes
- No

What was the surgery?

Surgery Description

When was it?

Month, Year

When was your last period?

- Less than 1 month ago
- 1-2 months ago
- 2-3 months ago
- 3-6 months ago
- 6-12 months ago
- 1-2 years ago
- 2-5 years ago
- 5-10 years ago
- 10+ years ago

Tell us more about your last period. (select all that apply)

- My period is light and spotting
- My cycles are shorter
- I’m skipping cycles
- Heavier than normal
- My period stopped after my hysterectomy
- My period stopped after placement of my hormonal IUD
- My period stopped after endometrial ablation
- None of these.

Are you pregnant or trying to become pregnant?

- Yes
- No
- I’m not sure

Are you using birth control?

- Yes
- No

What other kinds are you using? (select all that apply)

- Hormonal Pill
- Hormonal Patch
- Hormonal IUD
- Copper IUD
- Vaginal Ring
- Hormonal Shot
- Bilateral Tubal Ligation (removal of fallopian tubes)
- Other

Have you had a mammogram in the last two years?

- Yes
- No

When was your mammogram?

Month, Year

Did your mammogram show something abnormal?

- Yes
- No

What was the abnormality?

Benign (not cancerous)

Malignant (cancerous)

Undetermined

Please describe your mammogram results.

Are you currently using any kind of hormone therapy? This could include birth control, vaginal cream, or menopause hormone therapy.

- Yes
- No

Select all that apply.

- Pill
- Patch
- Transdermal spray, gel, cream
- Birth control pill
- Vaginal estrogen, vaginal DHEA, Osphena
- Pellet therapy
- Other

When was your last dose?

- 0-3 months ago
- 3-6 months ago
- Over 6 months ago

Tell us what went well for you or what didn’t.

Describe your most recent dose and what your experience was

Are you having any unexplained uterine or vaginal bleeding?

- Yes
- No

When did that start?

- 1-2 months ago
- 2-3 months ago
- 3-6 months ago
- 6-12 months ago
- 1-2 years ago
- 2-5 years ago
- 5-10 years ago
- 10+ years ago.

Please enter your most recent blood pressure. The top number is usually higher than the bottom number.

Systolic (mmHg)

- Less Than 90
- 90-139
- 140-149
- Greater than 150
- I don’t know

Diastolic (mmHg)

- Less than 50
- 50-79
- 80-89
- Greater than 90
- I don’t know

Have you been treated for any of the following medical conditions? Select all that apply

- Breast Cancer
- Uterine/Endometrial Cancer
- Ovarian Cancer
- Current Liver Disease
- Current Symptomatic Gallbladder Disease
- Current Asymptomatic (No Symptoms) Gallbladder Disease
- Systemic Lupus Erythematosus with Antiphospholipid Antibodies
- Coronary Heart Disease
- Heart Attack
- Stroke/TIA
- Deep Vein Thrombosis and/or Pulmonary Embolus
- Inherited Thrombophilia (e.g. Factor V Leiden)
- Current Atrial Fibrillation, Metastatic Melanoma
- Type 1 Diabetes
- Type 2 Diabetes
- Use of a wheelchair or limited mobility
- None of these

Please list all current and past medical conditions. Example: Colon cancer, high cholesterol, high triglyceride levels (hypertriglyceridemia), Roux-en-Y gastric bypass, biliopancreatic diversion surgery

Medical Conditions

I don’t have any medical conditions

Please list any prescription or over-the-counter medications and supplements you are taking. Please enter a comma after each item. For example, if you are taking Lipitor, write Lipitor to add this to your medication list. Example: Lipitor, multivitamin

Medications

I am not taking any medication or supplements

Please list your allergies. Please enter a comma after each allergy. For example if you are allergic to peanuts, write Peanuts, to add this allergy and repeat the process for each thing you are allergic to (including foods and medications).

Add a new allergy

I do not have any allergies

Do you use tobacco products?

- Yes
- No

What kinds of tobacco products do you use?

- Cigarettes
- e-Cigarettes/Vapes
- Other

How frequently do you use tobacco?

- One pack a day
- A few times a day
- A few times a week
- A few times a month

Is there anything else you’d like to share with your provider? Some members want to ensure their providers know about things like heart disease or family history of breast, ovarian, primary peritoneal, or uterine cancer. If you have a preferred treatment, such as Estradiol Patch, Estradiol Pill, non-hormonal options, or Vaginal Estradiol, you can share that here.
